# Efficacy and safety of a novel antiviral preparation in ICU-admitted patients with COVID-19: a phase III randomized controlled trial

**DOI:** 10.1101/2021.11.20.21266229

**Authors:** Hossein Faramarzi, Amirhossein Sahebkar, Ahmad Hosseinpour, Vahid Khaloo, Parisa Chamanpara, Mohammad Reza Heydari, Sajad Najafi, Fatemeh Fotoohi Khankahdany, Ahmad Movahedpour

## Abstract

**Introduction:** Despite an increasing number of studies, there is as yet no efficient antiviral treatment developed for the disease. In this clinical trial, we examined the efficacy of a novel herbal antiviral preparation comprising *Zataria multiflora Boiss, Glycyrrhiza glabra, Cinnamomum Vermont, Allium sativuml*, and *Syzygium aromaticum* in critically ill patients with COVID-19 patients.

**Methods:** A total number of 120 ICU-admitted patients requiring pulmonary support with a diagnosis of COVID-19 pneumonia were recruited to the trial. Participants were equally randomized to receive either the novel antiviral preparation sublingually, for up to two consecutive weeks or till discharge, or normal saline as the matching placebo. Clinical and laboratory parameters as well as survival rates were compared between the two groups at the study end.

**Results:** The cumulative incidence of death throughout the study period was 8.33% in the medication group and 60% in the placebo group (risk ratio: 0.14; 95% confidence interval [CI], 0.05 to 0.32; P<0.001). Survival rates were significantly higher in the treatment group. Additionally, on day 7, several laboratory factors including white blood cells (WBCs) count, C-reactive protein (CRP), and SpO_2_ were improved in patients treated with the novel antiviral preparation compared with the placebo group.

**Conclusion:** The novel antiviral preparation tested in this trial significantly improved the survival rate and reduced mortality in critically ill patients with COVID-19. Thus, this preparation might be suggested as a potentially promising COVID-19 treatment.

Funded by Shimi Teb Salamat Co., Shiraz, Iran, and registered on the Iranian registry of clinical trials (registration No. IRCT20200509047373N2).

## Introduction

The first cases of novel coronavirus disease (COVID-19) pandemic was initially reported in Wuhan city in the Hubei province of China during late December 2019 (1). By November 5, 2021, nearly a quarter billion people have been identified to be infected with the causative virus SARS-CoV-2 globally and the total worldwide death toll has exceeded 5 million (2). SARS-CoV-2 is an RNA virus, which attaches to the angiotensin-converting enzyme 2 (ACE2) receptor on the target cells *via* its spike (S) protein. Following entry into the host cells, the virus is fused to endosomes and uncoated. Then, primary proteins are translated and viral single-stranded RNA (ssRNA) genome is replicated. At the last steps, virions are assembled and released from the infected cells (3). SARS-CoV-2 is known to be highly transmitted through respiratory fluids including droplets and aerosol particles with higher transmission rates in asymptomatic infected individuals, which has made virus spread more prevalent and control of the disease more challenging (4). COVID-19 also demonstrates higher morbidity among affected patients with a broader age range and despite lower proportion of pediatric patients, severe disease is reported for COVID-19-affected children (5). The majority of COVID-19 patients recover from the disease without receiving medical care or hospitalization and receiving supportive treatments, while a proportion develop severe grades of the disease requiring mechanical ventilation and hospitalization in the intensive care unit (ICU). The highest mortality rates among COVID-19 cases have been reported in these critically ill patients as high as 50% to 67% (6, 7). These patients develop severe complication of COVID-19 like acute respiratory distress syndrome (ARDS), which along with extrapulmonary conditions are responsible for poor outcomes (8). Prolonged ICU hospitalization and invasive mechanical ventilation cause pulmonary infections leading to septic shock and multi-organ failure as the immediate contributing conditions to majority of deaths (9). Thus, management of ICU-hospitalized patients has faced the most challenges since the pandemic emergence. Globally, after development of highly efficient vaccines for SARS-CoV-2, mass vaccination has greatly helped preventing severe COVID-19 cases, hospital and ICU admissions, and total mortalities (10-13). Theoretically, several steps of viral life cycle and SARS-CoV-2-encoded proteins (e.g., viral proteases) could be inhibited to block the virus pathogenesis. However, despite a large number of studies, an efficient prophylactic or therapeutic treatment is yet to be discovered. Although several drugs such as corticosteroids, hydroxychloroquine lopinavir-ritonavir and remdesivir have been suggested as potential treatments, the findings of randomized controlled trials have been equivocal or suggested futility of some of these agents on the recovery of COVID-19 patients or decreasing the mortality rates (14-16). Considering the paucity of effective therapeutic agents, especially in critically ill patients, this study examined the efficacy of a herbal antiviral preparation on clinical symptoms, paraclinical parameters, and survival rates of ICU-admitted patients in a phase III clinical trial. This novel preparation is composed of several herbal ingredients including *Zataria multiflora Boiss, Glycyrrhiza glabra, Cinnamomum Vermont, Allium sativuml*, and *Syzygium aromaticum*. The preparation has previously shown antiviral activity in cell experiments and the current study is the first clinical experiment to investigate its efficacy in patients with COVID-19. Since the mortality among ICU-admitted patients is significantly higher due to underlying conditions and medications, we set out to evaluate the efficacy of antiviral preparation in critically ill COVID-19 patients. The results of this study can be extended to outpatients, who have less pulmonary involvement and less co-morbidities and mortality.

## Methods

### Trial Design and Participants

COVID-19 patients hospitalized in an ICU department of Shiraz University of Medical Sciences, Shiraz, Iran, were enrolled in a randomized, double-blind, placebo-controlled phase III clinical trial conducted during March 21-June 19,2021. The trial was registered on the Iranian registry of clinical trials (https://www.irct.ir) (registration No. IRCT20200509047373N2). The trial site was at Ali Asghar hospital, as one of the ICU centers specialized for caring COVID-19 patients in Shiraz city, to enroll the eligible subjects. ICU-admitted patients were included in the trial with several eligibility criteria including having confirmed diagnosis of COVID-19, high pulmonary involvement in imaging, requirement to respiratory support but not mechanical ventilation (however, they may require mechanical ventilation during trial), aged 15 years or older, filled informed consent, and not participated in other clinical trials. Pregnant women and patients younger than 15 years were excluded from the study. COVID-19 definite diagnosis was made by the infectious diseases specialist according to the national COVID-19 committee’s guidelines for diagnosis and management of COVID-19 patients *via* SARS-CoV-2 reverse transcriptase polymerase chain reaction (RT-PCR) positive results or through characteristic radiographic presentations in chest computerized tomography (CT) or radiographic imaging. Written informed consent was obtained from all patients or their legal representatives. The research was approved by the Ethics Committee of the Shiraz University of Medical Sciences (registration No. IR.SUMS.REC.1399.1367). Eligible patients were randomly assigned in a 1:1 ratio to either medication or placebo group.

### Sample Size

No prior data was available on the efficacy of the antiviral medicine in the management of COVID-19, so we performed a pilot study with 30 participants (15 per group) with the same protocol on similar patients and recorded the death rate as the preliminary data on the clinical efficacy of ROJA. According to the results, 3 (20%) of the participants in the ROJA group and 7 (46.7%) of those in the placebo group would have an event (death through day 14). By considering 90% power to detect a between-group difference and the alpha level of 5%, we estimated a sample of 120 patients (60 per group).

### Randomization and Interventions

Randomization was conducted by the statistical analyst using the Website Randomization.com (http://www.randomization.com) and the permuted block randomization method with a block size of 4. Patients were enrolled by a health worker rather than the research team and the statistical analyst performed the patient allocation. Allocation was masked to all individuals including patients, physicians and statistical analysts. Both groups received COVID-19 local standard treatments including remdesivir and glucocorticoids according to the guidelines. Patients assigned to the medication group received the antiviral preparation. Each patient was administered 1 ml (containing 433 mg of active materials) antiviral preparation in drop form using dropper every 3 hours for up to two consecutive weeks. Moreover, patients who underwent mechanical ventilation received the medicine up to 14 days or till discharge. Subjects in the placebo group were given a matching placebo containing normal saline in the same packaging with similar appearance, volume, and dosing as the active compound to maintain blinding. Standard care was provided for all patients in both groups.

### Antiviral preparation Composition and dosing

The compound is a novel antiviral drug with approved antiviral properties in cell culture experiment (data not published) formulated of several active herbal ingredients. Plant materials and concentrations included Zataria multiflora Boiss (5mg), Glycyrrhiza glabra (4mg), Cinnamomum (7mg), Allium sativuml (3mg), and Syzygium aromaticum (3mg). All lant materials were purchased from a local botanist in Iran and quality approved by a plant classification specialist in the Department of Botany, Shiraz University of Medical Sciences, Shiraz, Iran. All of them were cleaned, dried, mechanical powder, extracted with 100% Deionized water evaporated with hot plate at 90°C to prepare the extract and then filtered. Plant materials powder was mixed separately in 100 ml of distilled water. These suspensions were filtered by What Mann’s paper No. 1 and then mixed using a magnetic stirrer. The final crude extract was placed in the refrigerator until it was ready for use. The extract was used in drops with final concentration of 433±5 mg/ml to feed sublingually. The sterility test was performed to check the bacterial contamination in each extract. Nutrient agar plates were used to check the sterility of the extract by incubation at 37°C for 24 hrs. Reverse phase chromatographic analyses were carried out under gradient conditions using C18 column (4.6 mm × 250 mm) packed with 5 μm diameter particles. The extract was prepared in HPLC grade solvent. Then, the sample was sonicated using ultrasonicator for 10 min. The extract was filtered and injected into the HPLC column using mobile phase of 30:70 (acetonitrile:0.1% phosphoric acid). The above procedures were all performed at Shimi Teb Salamat Co., Shiraz, Iran

### Procedures

Throughout the trial since days 0 to 14, patients were assessed for their clinical status. Any adverse effect or exacerbation in outcomes was recorded. Several clinical and paraclinical parameters were evaluated in both groups on days 0, 7 and 14. Vital signs including body temperature, blood pressure, respiratory rate, and blood oxygen saturation (SpO_2_), laboratory parameters such as hemoglobin (Hb) values, white blood cells (WBCs) count, and C-reactive protein (CRP) levels were measured. SARS-CoV RT-PCR was conducted at the first time point *via* standard mid-nasal sterile swab samples. Pulmonary involvement was investigated via chest CT-scans. Co-existing conditions were also compared between medication and placebo groups. No significant change was applied in the final protocol compared to the initial planned protocol.

### Outcomes

The main primary outcome was the COVID-19-associated deaths, which was assessed through day 14 after enrollment. Other outcomes were clinical status, and changes in vital signs and laboratory parameters. Any possible adverse effect causing discontinuation of the study and impacting progression of the disease, as well as clinical or laboratory parameters were all recorded. No change was applied in the outcomes after trial was commenced.

### Statistical Analysis

The retrieved data was exported to SPSS software, version 18.0 (SPSS Statistics). To compare the qualitative and quantitative variables, Chi-square, independent samples t-test, paired samples t-test, and analysis of covariance (ANCOVA) were used and also, for non-normal variables the non-parametric Mann-Whitney U test and Wilcoxon signed-rank test were employed. Binary logistic regression was also used to estimate the odds ratio (OR) of death. Survival analysis was conducted using the Kaplan-Meier curve and Cox regression.

## Results

A total of 120 patients were recruited to the study. Patients were all confirmed with COVID-19 and hospitalized to ICU department for receiving intensive care. Advanced pulmonary involvement and requirement to respiratory support via positive pressure non-invasive ventilation was reported for all patients. Among included participants, 68 patients (56.7%) were male and 52 patients (43.3%) were female. Mean age was 60.29*±*12.94 years with a range of 29-97 years (Table 1). Co-existing conditions, like cardiovascular disease, diabetes, or chronic respiratory disease was seen for 84 patients (70%) (table 1). 60 patients were randomly assigned to the medication group and an equal number assigned to the placebo group. All patients completed the study, either recovered or died, and no participant left the trial (figure 1). Table 1 shows some baseline information for both groups at day 0. Groups demonstrated homogenous assignment for majority of parameters, however, despite random assignment age mean was different between groups. Therefore, in order to adjust the age effect as a confounding variable we used analysis of covariance (ANCOVA) test to compare the groups on the 7th day.

**Table 1.**
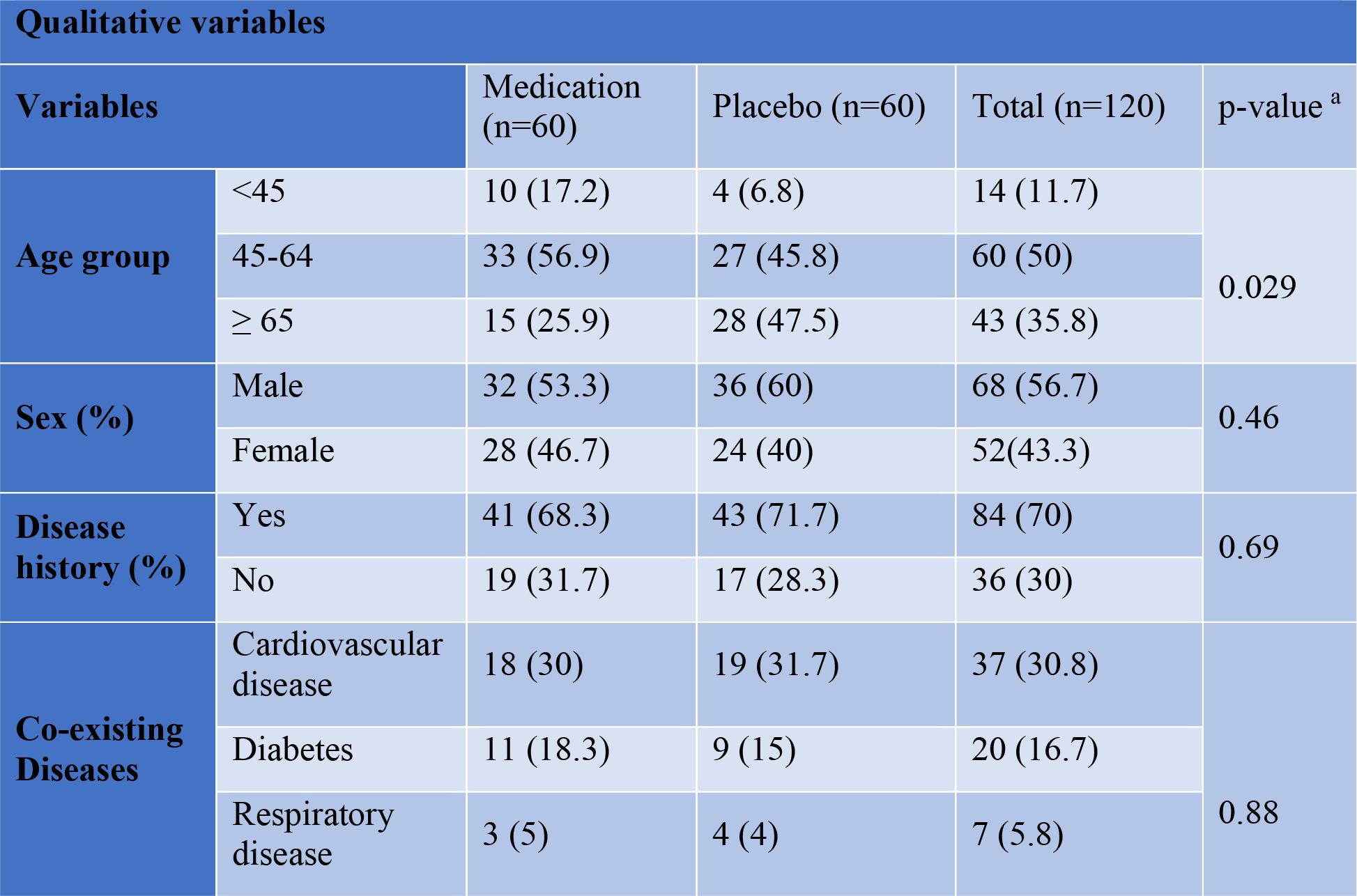
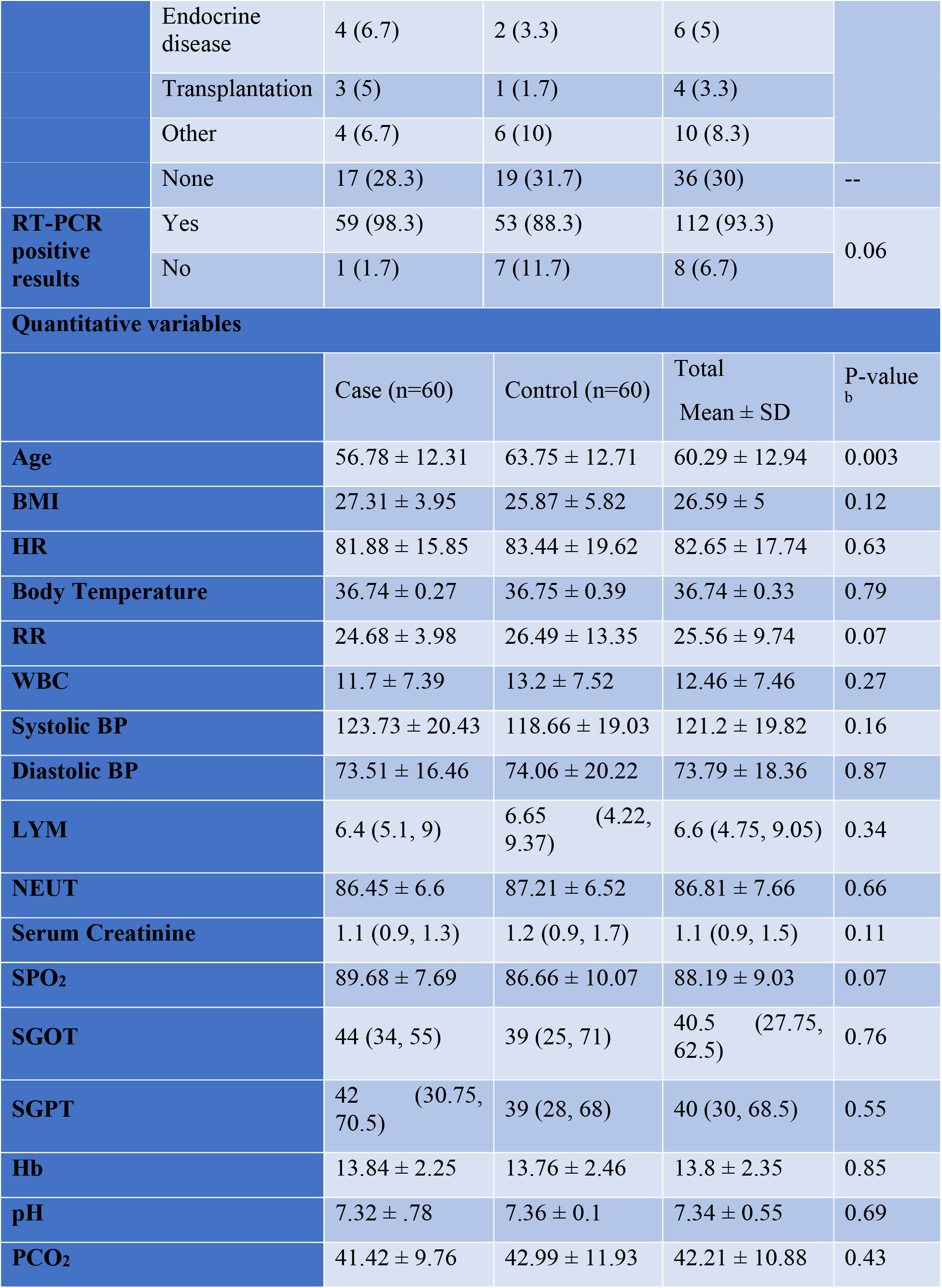
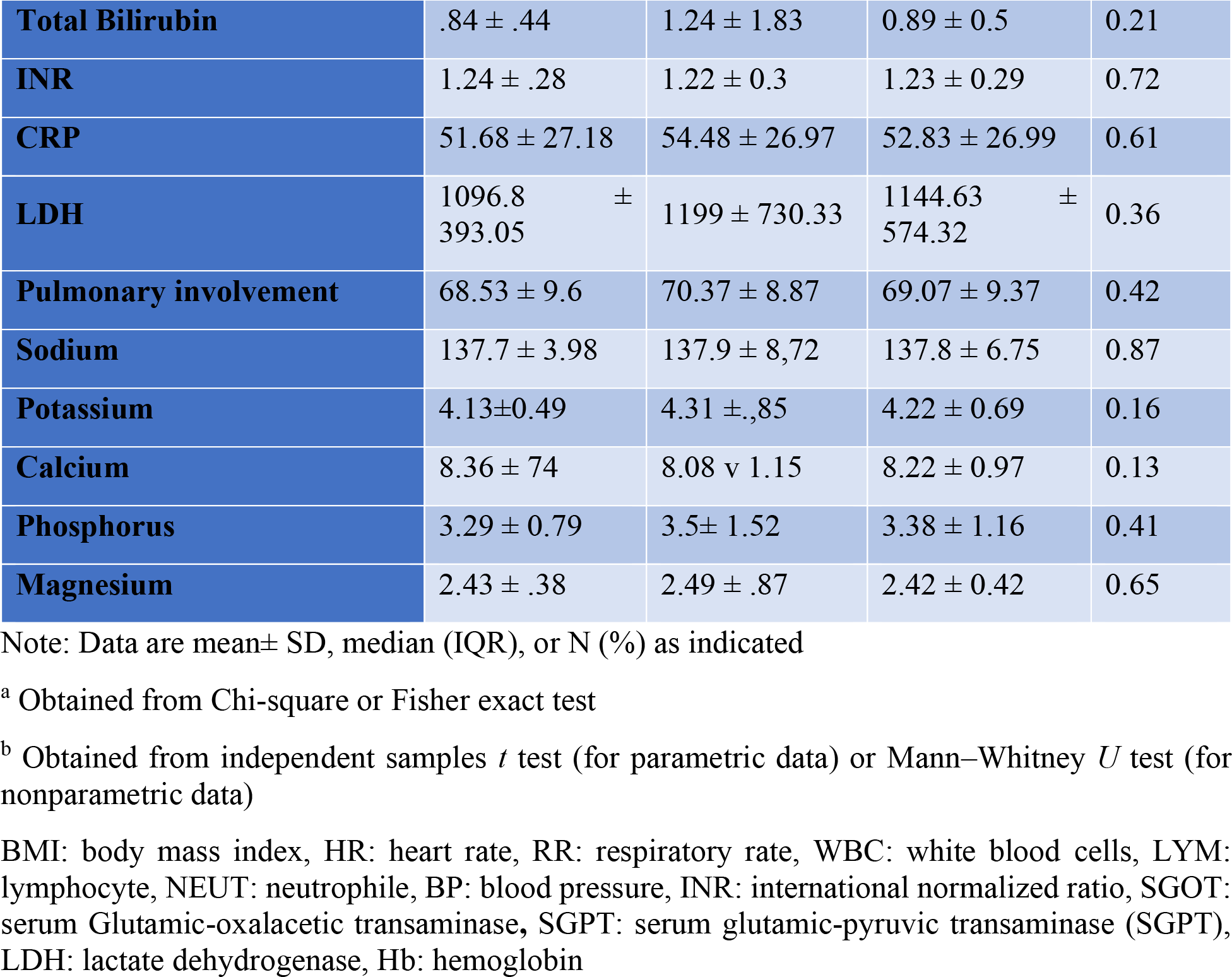
Baseline clinical and laboratory values for medication and placebo groups.

**Figure 1.**
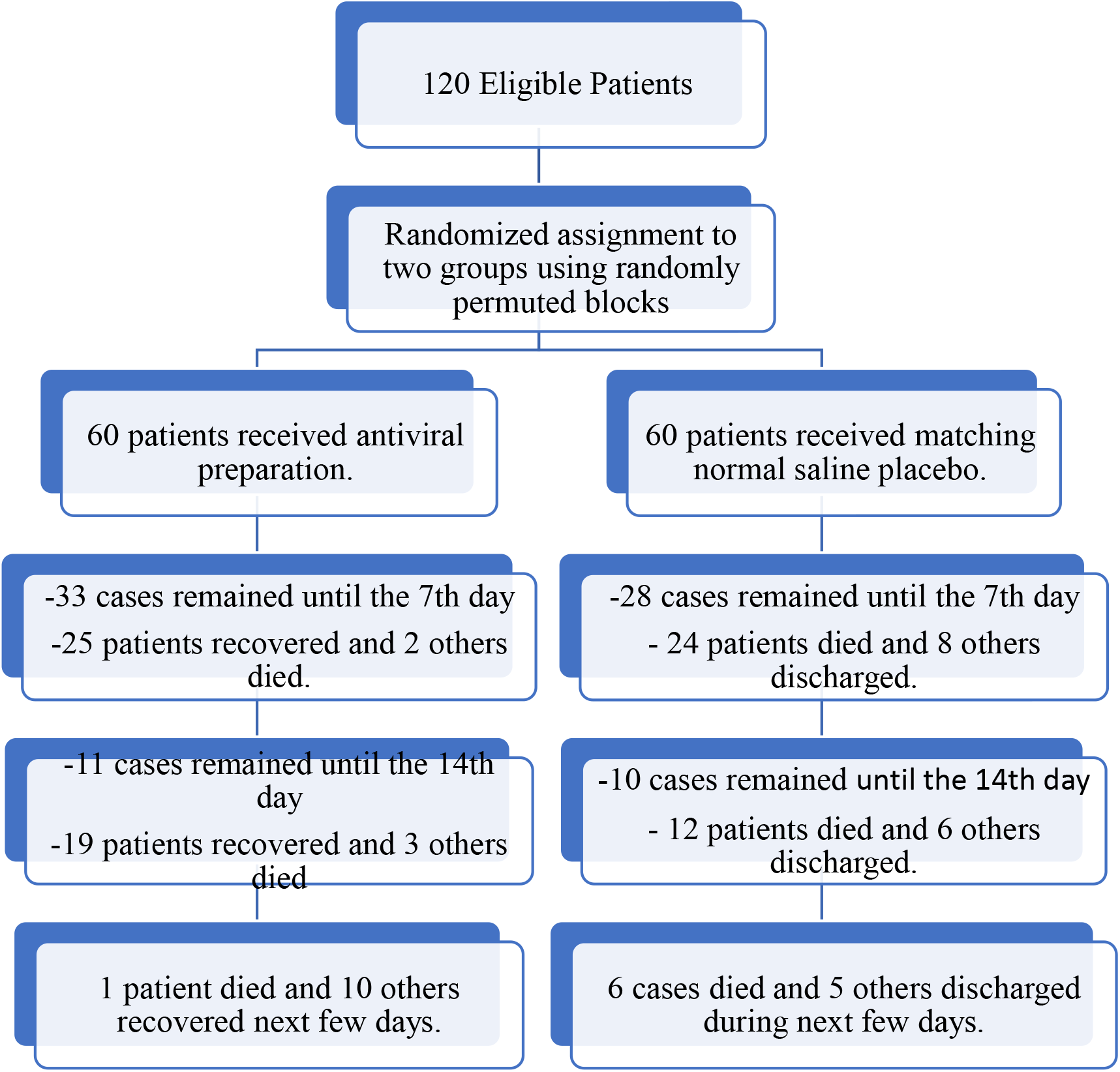
Flow diagram of trial.

### Primary Outcomes Mortality

Until day 14, death occurred in 5 patients (8.33%) in the antiviral preparation group, while in 36 patients (60%) of the control group, which means highly significant difference in mortality between medication and placebo groups. The probability of death for patients received the medication was 0.14 times of those for the placebo group (risk ratio: 0.14; 95% confidence interval [CI], 0.05 to 0.32; P<0.001). To determine the odds ratio of mortality, logistic regression analysis was conducted. The results demonstrated that the age-adjusted odds ratio of death for the placebo group compared to the antiviral preparation group was 14.63 (95% confidence interval [CI], 4.95 to 43.24; P<0.001). We also evaluated the effect of the intervention on median survival during 14 days as age-adjusted hazard ratio derived from Cox regression was 0.12 (95% confidence interval [CI], 0.04 to 0.31; P<0.001). The Kaplan-Meier curve along with Log Rang test also showed significant difference in the survival rate between both groups (figure 2). In fact, the patients received the antiviral preparation were predicted with higher survival compared to the placebo group.

**Figure 2.**
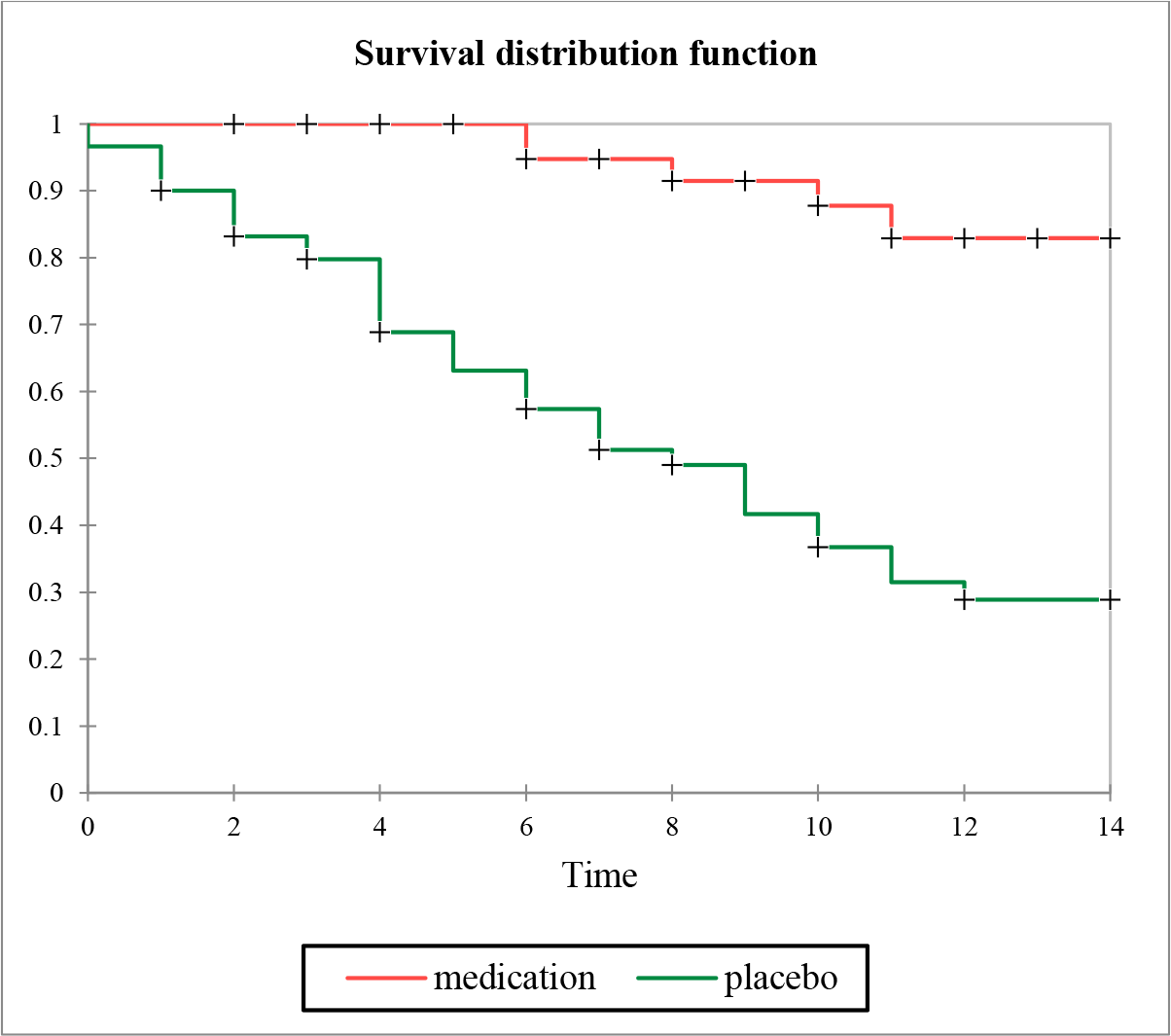
Survival analysis in Kaplan-Meier analysis shows decreased survival in placebo group compared to the control group

### Secondary Outcomes

Due to low sample size of the placebo group resulted from missed dead patients on the 14th day, improvement in several clinical and laboratory parameters of patients was evaluated through day 7 (Table 2). Some respiratory parameters like respiratory rate (RR) showed significant improvement in patients treated with the antiviral preparation on day 7 compared to the baseline measurement; however, the change was not significant compared to the placebo group. Several laboratory parameters like peripheral blood lymphocyte and neutrophile count, and total bilirubin showed improvement in the antiviral preparation-treated patients, while lymphocyte count and Serum glutamic pyruvic transaminase (SGPT) exacerbated in placebo group. Overall, white blood cells (WBCs) count, CRP, and calcium were improved in antiviral preparation-treated patients compared to the placebo group. Importantly, SpO_2_ demonstrated highly significant improvement in the medication participants compared to the placebo group. No adverse effect was reported for the patients in association with the treatment.

**Table 2.** Clinical and laboratory petameters post intervention.

| **Variables** | **groups** | **Day 0** | **Day 7** | **p-value <sup>a</sup>**<br>**(Within group)** | **p-value <sup>b</sup>**<br>**(Between group)** |
| --- | --- | --- | --- | --- | --- |
| **HR** | Medication | 82.03 ± 17.84 | 85.2 ± 13.84 | 0.28 | 0.05 |
|  | Placebo | 87.44 ± 16.45 | 93.32 ± 13.67 | 0.13 |  |
| **Body Temperature** | Medication | 36.71 ± 0.28 | 36.7 ± 0.32 | 0.86 | 0.32 |
|  | Placebo | 36.75 ± 0.24 | 36.8 ± 0.36 | 0.41 |  |
| **RR** | Medication | 24.93 ± 3.55 | 22.71 ± 3.26 | 0.001 | 0.53 |
|  | Placebo | 23.8 ± 5.49 | 23.07 ± 2.93 | 0.52 |  |
| **WBC** | Medication | 12.4 ± 9.49 | 12.2 ± 4.84 | 0.92 | 0.05 |
|  | Placebo | 12.45 ± 6.33 | 15.56 ± 7.55 | 0.08 |  |
| **Systolic BP** | Medication | 125.69 ± 21.54 | 124.54 ± 15.48 | 0.66 | 0.18 |
|  | Placebo | 118.48 ± 18.77 | 116.76 ± 14.75 | 0.69 |  |
| **Diastolic BP** | Medication | 74.21 ± 12.83 | 75.03 ± 11.41 | 0.73 | 0.69 |
|  | Placebo | 77.11 ± 28.75 | 73.64 ± 13.7 | 0.56 |  |
| **LYM** | Medication | 9.12 ± 10.31<br>6.15 (5.15, 9) | 12.49 ± 12.14<br>8.7 (5.95, 16.37) | 0.007 | 0.31 |
|  | Placebo | 7.14 ± 4.01<br>5.6 (4.6, 9) | 8.9 ± 5.11<br>7.1 (5.5, 12.85) | 0.04 |  |
| **NEUT** | Medication | 85.64 ± 10.95 | 79.42 ± 13.69 | 0.003 | 0.28 |
|  | Placebo | 87.9 ± 5.42 | 84.51 ± 7.73 | 0.03 |  |
| **Serum Creatinine** | Medication | 1.1 (0.9, 1.4) | 1.2 (0.9, 1.4) | 0.93 | 0.58 |
|  | Placebo | 1.2 (0.9, 1.45) | 1.05 (0.8, 1.3) | 0.21 |  |
| **SpO<sub>2</sub>** | Medication | 89.68 ± 7.69 | 92.7 ± 3.39 | 0.005 | <0.001 |
|  | Placebo | 87.54 ± 10.23 | 83.74 ± 13.49 | 0.07 |  |
| **SGOT** | Medication | 44 (34, 55) | 43 (30, 54) | 0.35 | 0.32 |
|  | Placebo | 38.5 (24.75, 65) | 39 (23.75, 90) | 0.3 |  |
| **SGPT** | Medication | 37 (29.75, 58.25) | 57 (35.25, 75) | 0.09 | 0.82 |
|  | Placebo | 36.5 (29.25, 76) | 60.5 (42, 89.5) | 0.03 |  |
| **Hb** | Medication | 13.52 ± 2.4 | 13.38 ± 2.05 | 0.61 | 0.32 |
|  | Placebo | 14 ± 2.46 | 13.28 ± 2.88 | 0.1 |  |
| **pH** | Medication | 7.22 ± 1.06 | 7.23 ± 1.06 | 0.6 | 0.07 |
|  | Placebo | 7.37 ± 0.1 | 7.15 ± 0.78 | 0.12 |  |
| **PCO<sub>2</sub>** | Medication | 41.44 ± 10.71 | 43 ± 9.21 | 0.36 | 0.44 |
|  | Placebo | 40.75 ± 9.3 | 45.88 ± 14.02 | 0.06 |  |
| **Total Bilirubin** | Medication | 0.9 ± 0.55 | 1.16 ± 0.7 | 0.02 | 0.81 |
|  | Placebo | 0.87 ± 0.33 | 1.1 ± 0.42 | 0.08 |  |
| **INR** | Medication | 1.23 ± 0.23 | 1.14 ± 0.23 | 0.1 | 0.17 |
|  | Placebo | 1.15 ± 0.18 | 1.22 ± 0.29 | 0.29 |  |
| **CRP** | Medication | 53 (27, 72.75) | 15 (5.5, 37) | <0.001 | 0.01 |
|  | Placebo | 58 (50, 73) | 60 (23, 76) | 0.47 |  |
| **LDH** | Medication | 1161.73 ± 440.7 | 1015.56 ± 451.82 | 0.06 | 0.99 |
|  | Placebo | 1129.76 ± 447.36 | 999.82 ± 447.36 | 0.44 |  |
| **Sodium** | Medication | 137.6 ± 4.19 | 137.18 ± 3.8 | 0.62 | 0.83 |
|  | Placebo | 139.27 ± 12.39 | 138.15 ± 6.39 | 0.58 |  |
| **Potassium** | Medication | 4.21 ± 0.53 | 4.27 ± 0.48 | 0.61 | 0.42 |
|  | Placebo | 4.28 ± 0.81 | 4.11 ± 0.68 | 0.49 |  |
| **Calcium** | Medication | 8.18 ± 0.83 | 8.59 ± 0.65 | 0.02 | 0.031 |
|  | Placebo | 8.08 ± 0.99 | 8.25 ± 0.75 | 0.53 |  |
| **Phosphorus** | Medication | 3.44 ± 0.83 | 3.52 ± 1.15 | 0.74 | 0.81 |
|  | Placebo | 3.39 ± 1.48 | 3.32 ± 0.99 | 0.82 |  |
| **Magnesium** | Medication | 2.36 ± 0.33 | 2.31 ± 0.28 | 0.44 | 0.54 |
|  | Placebo | 2.45 ± 0.42 | 2.4 ± 0.49 | 0.7 |  |

Note: Data are mean ± SD or median (IQR) as indicated
<sup>a</sup>: Obtained from pair sample *t* test (for parametric data) or Wilcoxon (for nonparametric data)
<sup>b</sup>: Obtained from Analysis of Covariance test, by adjusting age and baseline value

## Discussion

The world has been facing the COVID-19 pandemic since late 2019, which has affected millions of lives and threatened billion others. Little is known about the pathophysiological details of SARS-CoV-2 virus, while it affected both sexes, wide age range (although with lower pathogenesis in children compared to more severe disease in elderly), races, and particularly individuals with comorbidities like diabetes, hypertension, and underlying pulmonary diseases in all continents. A majority of COVID-19 cases are asymptomatic, recover from mild illness spontaneously or after receiving supportive therapies. However, millions of deaths have resulted from severe pulmonary involvement contributing to multi-organ failure. Highest rates of mortality among hospitalized patients (up to 88%) have been reported in ICU-admitted cases, which require intensive pulmonary support due to advanced lung involvement and co-morbidities (17).

Taking less than one year, SARS-CoV-2 vaccines have been developed most quickly compared to any other vaccine in the history (18). To date, several vaccines have received emergency or final approval for clinical use, a number of others are in the pipeline, and nearly 4 billion people (more than half of the global population) have received at least one dose worldwide (19). Globally, these vaccines have shown robust effects in the prevention of severe disease and subsequently reduced COVID-19-associated mortality, particularly among older and susceptible individuals (20-22). However, new SARS-CoV-2 infections, although mainly milder, have been reported to occur in a number of vaccinated people (23). Moreover, to achieve high protection in a society, the majority of the population are required to get vaccinated since unvaccinated individuals may drive further outbreaks (24). This may take more extended time, investment, and comprehensive policies to support mass vaccinations in addition to requirements for global equal distribution. Additionally, high effectiveness of SARS-CoV-2 vaccines against novel viral variants resulted from frequent mutations is a place of doubt requires further investigations and may need optimizations (25). Due to lack of definite preventive or therapeutic approaches, conventional interventions like face masks, and social distancing are still recommended as non-pharmaceutical interventions particularly in settings with high possibility of transmission in addition to vaccination for breaking the virus cycle until its eradication (26).

Hitherto, no efficient therapy has been developed for COVID-19 patients and several suggested treatments like convalescent plasma taken from already infected patients, and repurposed drugs like chloroquine, hydroxychloroquine, ribavirin, lopinavir-ritonavir, favipiravir, and ivermectin have failed in several clinical trials to demonstrate significant impact on the course of the disease (27-29). Although studies have shown reduced hospitalization, recovery time and mortality with the use of a number of drugs like antiviral remdesivir (during the early phase of disease), interferons, and dexamethasone (30); these agents could not be considered as definite treatments for COVID-19 as their efficacies in other studies have been questioned and several drug agencies have not approved their clinical use unlike others.

In this study, we have introduced a novel herbal antiviral preparation with significant effects in critically ill patients with COVID-19. In this randomized, double-blind, placebo-controlled phase III trial, we showed the efficacy and safety of the antiviral preparation in ICU-hospitalized patients. In our previous study, we showed that the antiviral preparation inhibited viral load of RNA viruses (data not published yet). Our results demonstrated 91.67% recovery of patients after up to two weeks of treatment, and 8.33% mortality rate in the antiviral preparation-treated patients compared with 40% recovery and 60% mortality in the placebo group. These numbers unveiled a highly significant impact of the antiviral preparation in reducing COVID-19-associated deaths. COVID-19 patients in the control group, who received normal saline, had age-adjusted odds ratio (OR) of death of about 14.63 compared with those given the antiviral preparation. Survival analysis also demonstrated a significantly higher survival rate in the antiviral preparation-treated versus control patients. Additionally, clinical and laboratory parameters demonstrated improvements in the treatment group after 7 days compared with placebo-prescribed patients.

## Conclusion

The herbal antiviral preparation demonstrated high efficacy in the improvement of critically ill ICU-admitted patients with COVID-19. It significantly reduced mortality rate, and improved clinical and laboratory parameters compared to the placebo group. Taken together, the novel antiviral preparation can be considered as a potential therapeutic agent for SARS-CoV-2-infected patients not only for critically ill patients but also for outpatients. Further investigations are recommended to discover the therapeutic effects of the antiviral preparation in different subsets of COVID-19 patients along with mechanistic pharmacological studies.

## Data Availability

All data produced in the present study are available upon reasonable request to the authors.

http://www.irct.ir

## Acknowledgment

The authors acknowledge the health workers and officials at Ali Asghar Hospital as well as our colleagues at Shiraz Food and Drug Administration, Health Policy Research Center, Ethical Committee, Deputy of Treatment and Chancellor of Shiraz University of Medical Sciences who enabled us to perform successfully the study.

## Figure legends

**Figure 1.** A total number of 120 ICU-admitted COVID-19 patients were recruited to the study and randomly assigned to two group of medication and intervention. the antiviral preparation-treated patients demonstrated lower deaths compared to the control group on days 7, 14, and post-14.

Figure 1. Flowchart of the study

